# Movement-specific effects of transcranial magnetic stimulation over premotor and parietal cortices after stroke

**DOI:** 10.64898/2026.10.22.26365984

**Authors:** Golnaz Haddadshargh, Roberto M. de Freitas, Jennifer Mak, Amy Boos, Xiaoqi Fang, Jennifer L. Collinger, Gina McKernan, Liang Zhan, Fang Liu, George F. Wittenberg

**Author notes:** Correspondence: Golnaz Haddadshargh.

## Abstract

**Background:** Transcranial magnetic stimulation (TMS) has been proposed to support post-stroke motor recovery, but the response may depend on the task and motor-network status. We aimed to compare how premotor and parietal stimulation affects performance across reach directions with different movement demands, in preparation for therapeutic application of TMS.

**Methods:** 16 participants with chronic subcortical stroke performed planar reaches with the KINARM exoskeleton toward distal and proximal targets. Before movement, a 10-Hz triple-pulse TMS train was delivered over bilateral dorsal or ventral premotor cortex (PMd, PMv) or dorsomedial posterior parietal cortex (PPC) at subthreshold and suprathreshold intensities. The parietal midline was used as the control target. Participant-level bootstrap confidence intervals assessed changes in trajectory straightness and smoothness relative to the control target and differences between reach directions. We compared motor-network activity and connectivity using task-based and resting-state functional magnetic resonance imaging (fMRI) data from participants with stroke and neurologically intact controls. To explore response variability, we tested associations between TMS response and motor impairment, resting-state connectivity, and the change in task-evoked activity at the stimulated region from planning to movement.

**Results:** Suprathreshold PMd and PPC stimulation in both hemispheres improved trajectory straightness or smoothness during distal reaches relative to the control target. Suprathreshold contralesional PMd stimulation increased trajectory deviation and reduced smoothness during proximal reaches. Baseline performance during distal reaches was more closely related to motor impairment, and contralesional PMd effects were more favorable during distal than proximal reaches across all kinematic measures. Participants with stroke showed lower bilateral PMv activation during movement and stronger interhemispheric PPC connectivity. Only the change in task-evoked activity at the stimulated region from planning to movement was related to TMS response. Smaller changes relative to controls were associated with more favorable kinematic effects during distal reaches.

**Conclusions:** Movements more affected after stroke were the most responsive to this type of stimulation. The response to TMS was also associated with task-evoked activity differences at the stimulated region between movement planning and execution. Neuromodulation may therefore be most effective when designed around both the pathophysiology of the cortical target and the type of movement.

**Trial registration:** ClinicalTrials.gov, NCT04286516. Registered 21 January 2020.

## 1. Background

Persistent upper-limb impairment after stroke has motivated efforts to support motor recovery using noninvasive brain stimulation [1], but responses vary substantially with lesion anatomy, residual motor capacity, stimulation parameters, and the physiological state of the motor system [2,3]. Task demands and movement phase may also influence the response by shaping interactions among premotor, parietal, and primary motor regions [4]. Studies in individuals with stroke show distinct behavioral effects of contralesional motor and parietal stimulation across tapping, pointing, and reach-to-grasp tasks [5,6]. Even within a single reaching task, the effects of transcranial direct current stimulation (tDCS) over primary motor cortex (M1) on reaching distance varied with shoulder-abduction load in individuals with chronic stroke [7].

In neurologically intact adults, the selected reach goal is encoded before movement by the superior parietal lobule (SPL) within the posterior parietal cortex (PPC) in both hemispheres and by dorsal premotor cortex (PMd) contralateral to the moving arm [8]. SPL and PMd contribute differently to trajectory planning. SPL represents direct target-oriented trajectories, whereas PMd carries information about both direct and more complex paths [9]. During a wrist task, transcranial magnetic stimulation (TMS) over the medial intraparietal area within PPC disrupted movement direction, and TMS over contralateral PMd disrupted movement amplitude [10]. Recordings during reach-to-grasp movements in macaques also suggest that PMd and ventral premotor cortex (PMv) have different roles in movement planning, as instruction cues evoke stronger activity in PMv than in PMd [11].

After stroke, the relative contribution of each hemisphere depends partly on the integrity of the ipsilesional motor system. When corticospinal integrity is relatively preserved, motor control may be supported through ipsilesional premotor cortex, whereas greater corticospinal damage may increase the contribution of contralesional M1 through the remaining cortico-cortical pathways [12]. Responses to inhibiting contralesional M1 and facilitating contralesional PMd likewise varied with impairment severity and corticospinal damage [13]. Contralesional SPL may also contribute to recovered motor control, as TMS over this region disrupted the timing and accuracy of complex finger movements after subcortical stroke [14]. Motor pathway integrity may therefore be an important source of variability in the response to stimulation after stroke.

Designing stimulation for a specific movement after stroke also requires knowing whether the effect of perturbing a given cortical region remains consistent when features such as reach direction change. Our previous study showed that TMS over non-primary motor areas altered trajectory straightness and smoothness in neurologically intact adults [15]. These effects appeared similar across reach directions and were therefore analyzed together. After stroke, however, changes in the pathways available for motor control and the relative contributions of the two hemispheres may make stimulation effects more sensitive to movement demands. We therefore tested how the effects of premotor and parietal stimulation varied between reach directions requiring different patterns of shoulder and elbow motion.

Here, participants with chronic subcortical stroke performed reaches toward distal and proximal targets using their affected arm while a three-pulse, 10-Hz TMS train was delivered before movement over bilateral PMd, PMv, and dorsomedial SPL within PPC. Changes in trajectory straightness and smoothness were quantified separately for the two reach directions. Using functional magnetic resonance imaging (fMRI), we compared task-evoked activity and resting-state functional connectivity between the stroke and neurologically intact control groups to identify differences in regional activity and connectivity among premotor, parietal, and primary motor regions after stroke. To understand the varied effects of stimulation, we used correlational analyses to relate TMS response to motor impairment severity, resting-state connectivity patterns identified within the stroke group, and the change in task-evoked activity at the stimulated region from planning to movement.

## 2. Methods

### 2.1 Participants

Study protocols were approved by the Institutional Review Boards of the University of Pittsburgh (study #19080097) and Veterans Affairs (study #1617353) and registered at ClinicalTrials.gov (NCT04286516). Participants were recruited through the Veterans Affairs Pittsburgh Healthcare System and UPMC Healthcare Systems and provided written informed consent before enrollment.

22 potential participants with stroke were consented. One was determined to be ineligible, and one withdrew before data collection, leaving 20 participants with stroke who contributed data to at least one analysis. Their individual characteristics and data availability are presented in Table 1. 26 neurologically intact control participants were also enrolled (16 females, 10 males; 24 right-hand dominant, 2 left-hand dominant; mean age ± standard deviation: 63.6 ± 6.8 years; age range: 47–79 years). This cohort and its associated MRI data were reported previously and were included here as the control group [15].

**Table 1.**
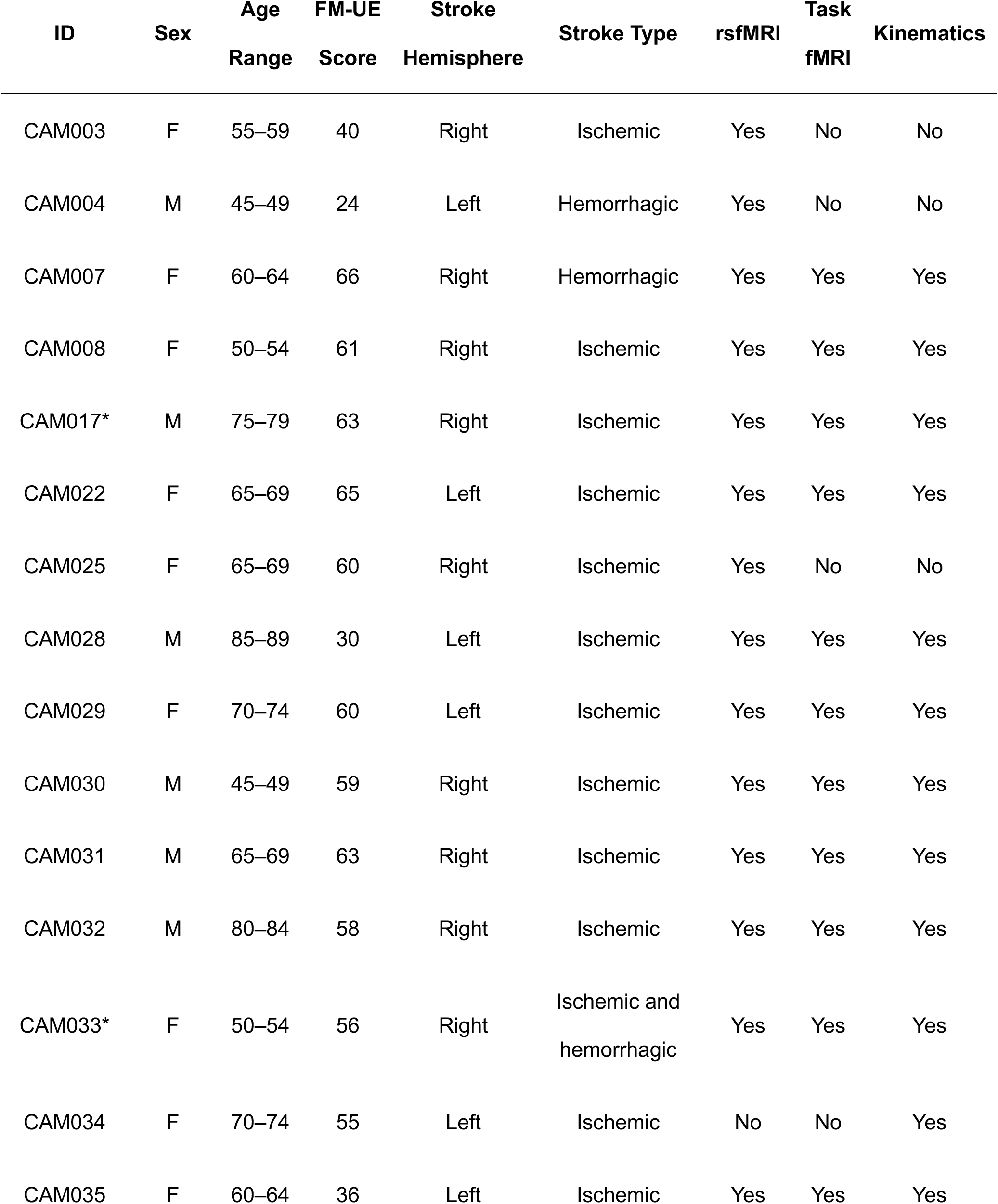
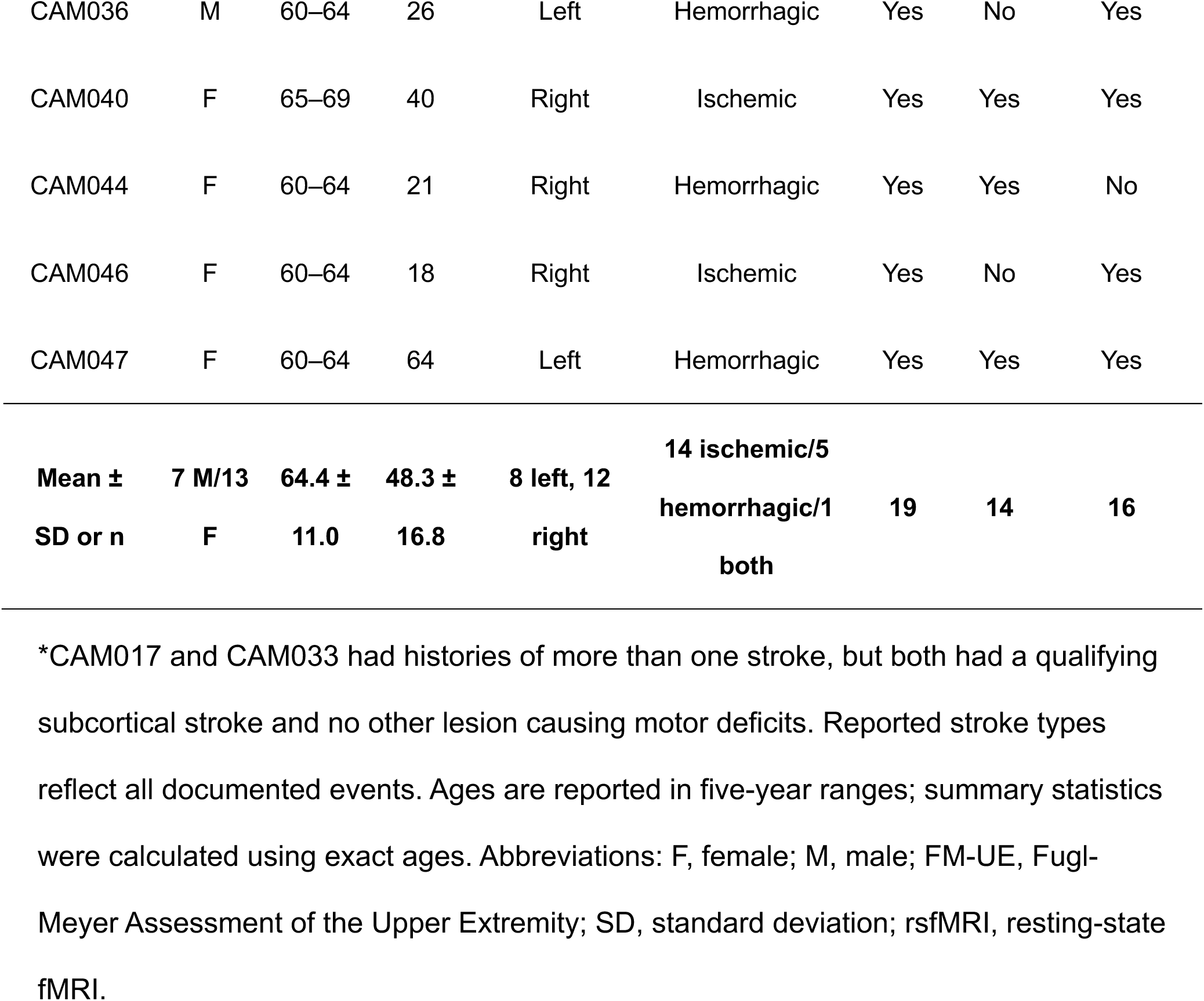
Participant characteristics and data availability.

Participants in both groups were eligible if they were 45–90 years old, medically stable, and able to speak English. Participants with stroke additionally required a clinically defined, unilateral hemiparetic stroke with alternative diagnoses excluded radiologically; a qualifying subcortical lesion; at least 6 months since stroke onset; and clinically assessed mild-to-moderate arm involvement with sufficient voluntary movement to perform the reaching task using the affected arm. Additional eligibility criteria for controls were adequate language and neurocognitive function to complete study training and testing.

Exclusion criteria for both groups included inability to provide informed consent or comply with study procedures; serious medical illness; contractures, orthopedic problems, visual loss, or other impairments that would interfere with testing; concurrent participation in another greater-than-minimal-risk study; contraindications to TMS or MRI; and pregnancy.

Resting-state fMRI comparisons included 19 participants with stroke and 25 controls, and task-based fMRI comparisons included 14 participants with stroke and 21 controls. The kinematic analysis included 16 participants with stroke. Differences in sample size were due to incomplete data from participants who did not complete all assessments or to data removal following quality-control and preprocessing procedures.

### 2.2 Experimental design

The study involved two experimental sessions on separate days. The MRI session included structural imaging, resting-state fMRI, and task-based fMRI, during which participants used a joystick to plan and execute visually cued movements. In the KINARM session, participants made planar reaches in two directions while TMS was delivered to six cortical targets and one control target. Stimulation site varied across blocks, and stimulation intensity varied across trials.

### 2.3 MRI procedure

#### 2.3.1 Acquisition parameters

MRI data were collected at the University of Pittsburgh Magnetic Resonance Research Center (RRID: SCR_025215) using a 3T Siemens Prisma scanner with a 64-channel head coil.

Structural images were collected to support anatomical registration, spatial normalization of the functional data, and individualization of TMS targets. High-resolution T1-weighted images were acquired using an MP-RAGE sequence with the following parameters: repetition time (TR) = 2300 ms, echo time (TE) = 2.9 ms, inversion time = 900 ms, flip angle = 9°, 1-mm isotropic voxels, and acquisition matrix = 256 × 256 × 192. T2-weighted images were acquired using a SPACE sequence (TR = 3200 ms, TE = 408 ms, 1-mm isotropic voxels, and acquisition matrix = 256 × 256 × 192).

Resting-state and task-based functional images were acquired using the same sequence parameters: multiband acceleration factor = 8, TR = 800 ms, TE = 37 ms, 2-mm isotropic voxels, and acquisition matrix = 104 × 104 × 72.

#### 2.3.2 fMRI Paradigms

Resting-state fMRI was acquired to compare connectivity among selected cortical regions involved in motor planning and movement between participants with stroke and controls. During acquisition, participants kept their eyes open and maintained fixation on a centrally displayed cross. A single run of 720 volumes was collected (9.6 min).

For task-based fMRI, participants performed a joystick-controlled task programmed in E-Prime 2.0 (Psychology Software Tools, Pittsburgh, PA, USA), preparing and executing horizontal movements toward targets presented on the left or right side of the screen. Participants with stroke used their affected arm, and control participants used their dominant arm.

The acquisition began with 6 s of central fixation. Each trial started with a variable fixation period of 2–4 s, followed by a 3-s directional cue indicating the upcoming movement direction. Participants then moved the joystick-controlled cursor toward the corresponding circular target. The 3-s movement period included approximately 1 s to move to the target, 1 s to maintain the target position, and 1 s to return to the starting point. Participants completed 60 trials, with leftward and rightward movements presented in randomized order (Figure 1).

**Figure 1.**
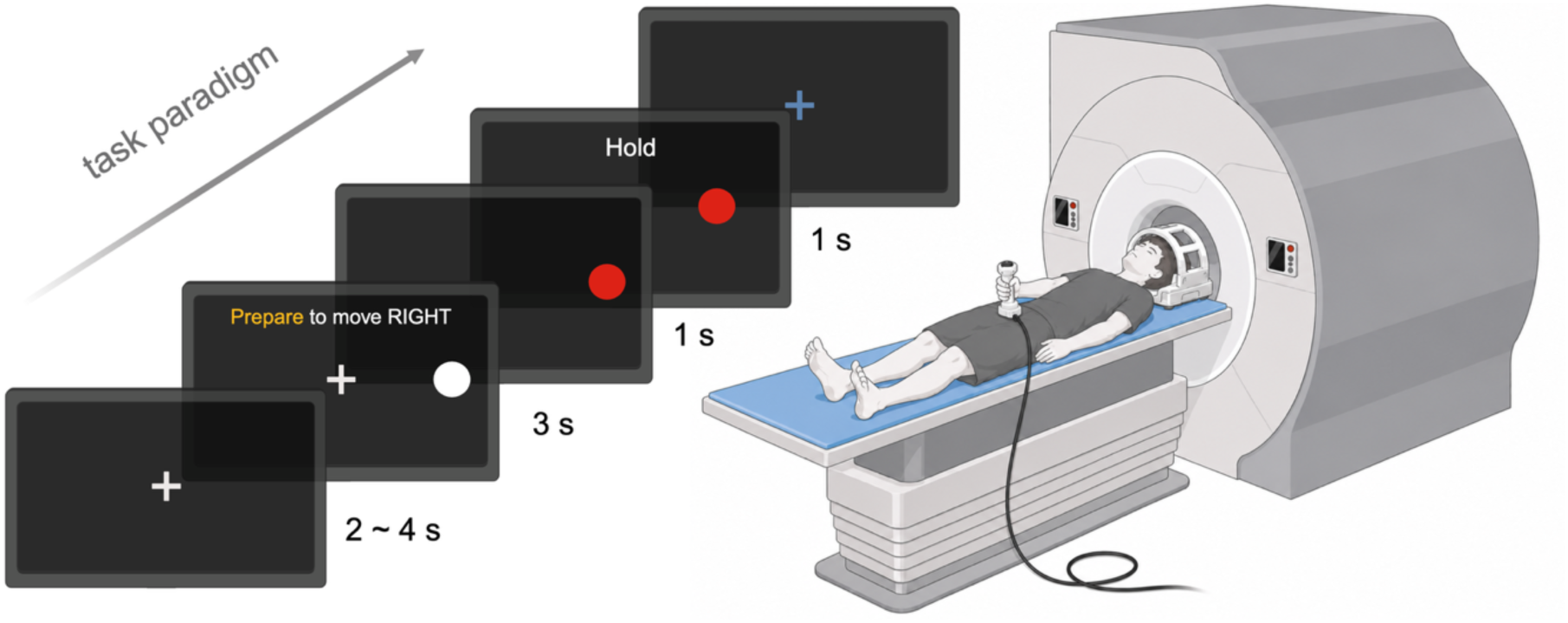
Task-based fMRI setup and task paradigm. Participants were positioned supine with their heads inside the head coil and used the tested hand to move a joystick held over the torso or thigh. The schematic shows an example rightward trial progressing from fixation to the directional cue, cursor movement, target hold, and return to the starting position, with the duration of each phase indicated.

### 2.4 KINARM task and TMS procedures

#### 2.4.1 Planar reaching paradigm

Participants performed a planar center-out reaching task using the KINARM Exoskeleton Robot (BKIN Technologies Ltd., Kingston, ON, Canada). They sat upright with their head supported by a headrest and both arms positioned in the robot’s arm supports. Reaching movements were performed with the affected arm (Figure 2a).

**Figure 2.**
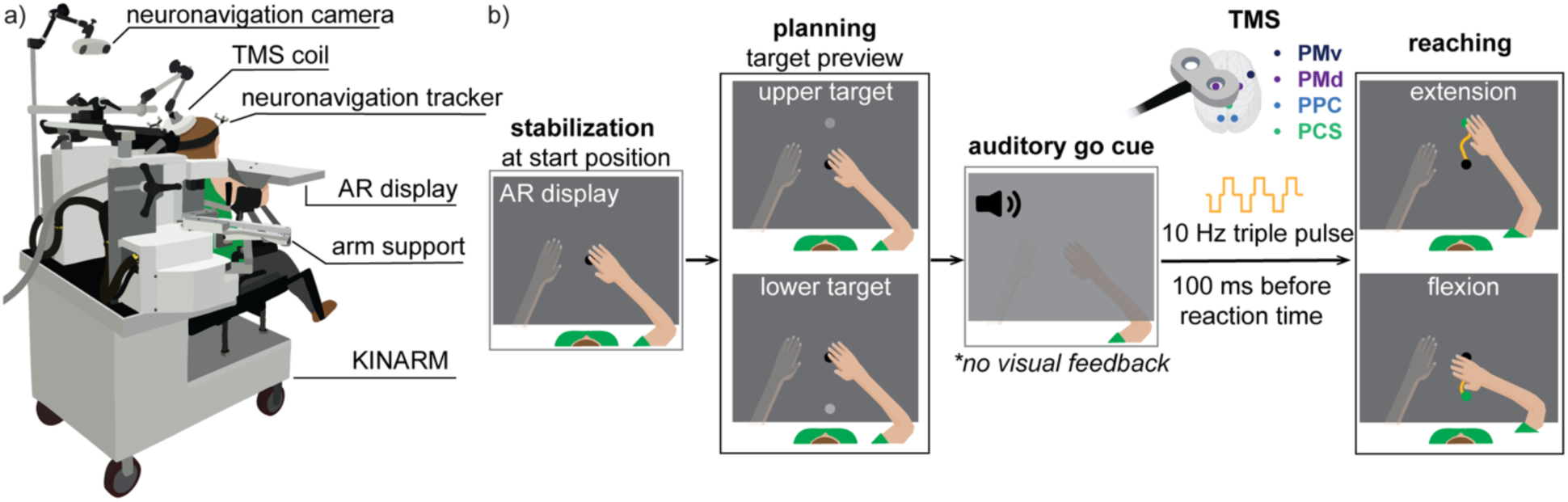
KINARM setup and planar reaching paradigm paired with TMS. (a) Experimental setup integrating the KINARM exoskeleton system with neuronavigated TMS. (b) Trial sequence for reaches toward the upper, distal and lower, proximal targets, referred to as extension and flexion reaches, respectively, based on the predominant elbow motion. A three-pulse, 10-Hz TMS train began 100 ms before the calculated reaction time. Abbreviations: PMv, ventral premotor cortex; PMd, dorsal premotor cortex; PPC, posterior parietal cortex; PCS, postcentral sulcus; TMS, transcranial magnetic stimulation.

Each trial began when participants stabilized their index fingertip at the central starting position for 1.5 s. A target then appeared at either an upper, distal location or a lower, proximal location. The target was initially positioned 11 cm from the starting point and adjusted when necessary to remain within the reachable workspace of the affected arm (range: 6–15 cm). During the 0.5–1.5-s planning period, the target and a white cursor representing the position of the index fingertip were visible on the display. At the auditory go cue, both the target and cursor disappeared. A blind positioned between the display and the arms prevented direct vision of the hands, so no visual feedback about fingertip position was available during the movement. Participants attempted to place their index fingertip at the remembered target location as quickly and accurately as possible (Figure 2b). Target direction was randomized across trials, with both directions presented equally often.

#### 2.4.2 TMS protocol

TMS was delivered using a PowerMAG PMD70-pCool double coil (Jali Medical, Framingham, MA, USA) supported by a flexible holder attached to the back of the KINARM. Stimulation targets included the bilateral PMd, PMv, and PPC, with the midline postcentral sulcus (PCS) as the control target. PMd targets were positioned anterior to the precentral sulcus and shifted slightly anteriorly to minimize direct stimulation of M1. PMv targets were identified using established anatomical landmarks, PPC targets were located within the dorsomedial SPL, and the PCS target approximated a medial postcentral site. The coordinates and the full selection rationale are reported in [15]. Target placement was guided by NeuroSynth meta-analysis maps [16].

Targets were individualized by registering the Montreal Neurological Institute (MNI152) atlas to each participant’s T1-weighted image using FMRIB’s Linear Image Registration Tool within the FMRIB Software Library (FSL; Oxford, UK) [17,18]. The transformed coordinates were imported into BrainSight (Rogue Research, Montréal, QC, Canada) and adjusted based on individual anatomy. Coil position was maintained using neuronavigation, with the coil held tangential to the scalp. The coil was oriented approximately 45° relative to the midsagittal line for PMd and PMv and along the anterior-posterior axis for PPC and PCS.

For motor-hotspot mapping and resting motor threshold (RMT) determination, a 3 × 3 stimulation grid with 10-mm spacing was defined in BrainSight and centered on the anatomical hand-knob region of the M1 contralateral to the unaffected arm. Surface electromyography (EMG) electrodes with integrated amplifiers (B&L Engineering, Santa Ana, CA, USA) were placed over the extensor digitorum communis (EDC) of the unaffected arm. Single-pulse TMS was delivered at each of the nine grid locations. The motor hotspot was identified as the location with the largest mean motor-evoked potential (MEP) amplitude and the most consistent responses of at least 50 μV peak to peak. RMT was then measured at the motor hotspot using the Motor Threshold Assessment Tool (MTAT) [19].

#### 2.4.3 Stimulation design

The KINARM task consisted of seven blocks, one for each stimulation target, with target order selected using a Latin-square design. Each block included 36 trials: 12 without stimulation, 12 with subthreshold stimulation at 80% of RMT, and 12 with suprathreshold stimulation at 120% of RMT, with 6 trials per movement direction in each condition. Stimulation conditions were randomized within each block. For RMT values greater than 83% of maximum stimulator output, suprathreshold stimulation was set at 100% of maximum output, while subthreshold stimulation remained at 80% of RMT.

Before the experimental blocks, participants completed at least 20 practice trials, which were used to calculate individual reaction time, defined as the interval between the auditory go cue and the point at which index-fingertip velocity exceeded 1 cm/s. Kinematic data were recorded at 1 kHz. TMS was delivered as a 10-Hz train of three pulses beginning 100 ms before each participant’s reaction time, targeting the transition from movement planning to execution.

### 2.5 Data analysis

#### 2.5.1 Kinematic data processing and outcome measures

To assess the effects of TMS on movement quality, reach trajectories recorded during the KINARM task were evaluated using straightness and smoothness metrics. Each trajectory was analyzed from 1 cm outside the starting point to 1 cm outside the target and then low-pass filtered using a zero-lag, sixth-order Butterworth filter with a 10-Hz cutoff. Straightness was quantified using path-to-length ratio (PLR) and trajectory deviation error (TDE), and smoothness was quantified using log-dimensionless jerk (LDJ) and spectral arc length (SAL) [20,21].

PLR represents the length of the observed trajectory relative to the shortest possible distance between the starting point and target:

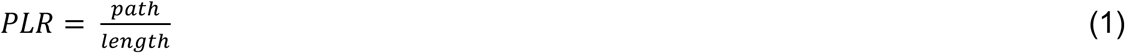

where *pat*ℎ is the length of the observed trajectory and *lengt*ℎ is the straight line connecting the starting point and target. TDE captures the cumulative spatial deviation of the observed trajectory from the straight-line path:

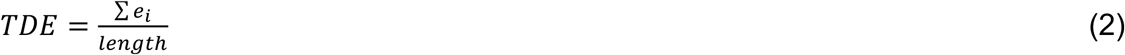

where *e_i_* is the minimum distance between the *i^th^* trajectory sample and the straight line connecting the start point to the end target. A perfectly straight reach has a PLR of 1 and a TDE of 0.

LDJ quantifies temporal smoothness using jerk (the rate of change of acceleration) normalized by movement duration and maximum velocity:

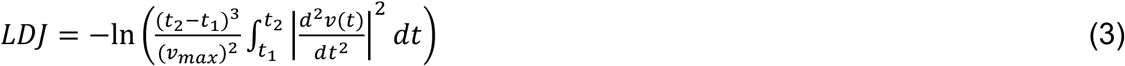

where *t*_1_ and *t*_2_ are the beginning and end of the analyzed trajectory, *v*(*t*) is hand velocity, and *v_max_* is maximum hand velocity. SAL quantifies smoothness in the frequency domain as the arc length of the normalized velocity spectrum:

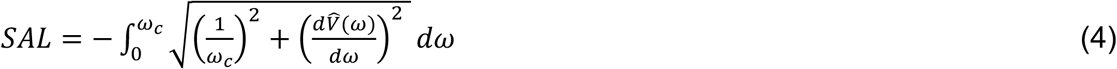

where *V*^ (*ω*) is the magnitude of the Fourier spectrum of *v*(*t*), normalized by the DC component *V*^ (0), and *ω* is the angular frequency over the range [0, *ω_c_*]. Consistent with previous studies, we adopted ω_c_ = 40π rad/s to capture the frequency range characteristic of human movement [20,22].

Before statistical analysis, LDJ and SAL were converted to absolute values, so that higher values indicated jerkier, less smooth trajectories. All four metrics were therefore oriented in the same direction, with higher values reflecting poorer movement quality.

#### 2.5.2 Analysis of TMS effects by movement direction

Reaches toward the upper, distal target primarily involved elbow extension and shoulder flexion, and reaches toward the lower, proximal target primarily involved elbow flexion and shoulder extension. Based on the predominant elbow motion in this task, we refer to the distal and proximal reach directions as extension and flexion, respectively, although both were multijoint reaches involving coordinated shoulder and elbow motion. The two directions were analyzed separately to identify direction-specific stimulation effects.

For each participant, kinematic metrics were averaged across trials for each combination of movement direction, stimulation target, and condition: no stimulation, subthreshold stimulation, or suprathreshold stimulation. For comparisons between a stimulation target and PCS, mean target distances were required to differ by less than 1 cm.

To account for differences in movement performance across participants, the effect at each PMd, PMv, and PPC target was expressed as the percentage change in each kinematic metric relative to PCS. Positive percentage changes indicated poorer performance relative to PCS, whereas negative changes indicated better performance. The same calculation was applied to no-stimulation trials to assess differences between the target and PCS blocks when no TMS pulses were delivered. For each metric, movement direction, stimulation target, and condition, participants were resampled with replacement 10,000 times to generate a 95% bootstrap confidence interval (CI) for the mean percentage change.

Extension and flexion were compared directly within participants when data were available for both directions for a given metric, stimulation target, and condition. For each participant, the flexion percentage change was subtracted from the extension percentage change, so negative values indicated a more favorable effect during extension and positive values a more favorable effect during flexion. The paired differences were bootstrapped as above. For both bootstrap analyses, the null hypothesis was rejected when the 95% CI excluded zero.

Movement direction may place different demands on residual motor capacity after stroke. We therefore assessed the relationship between impairment severity, measured using the Fugl-Meyer Assessment of the Upper Extremity (FM-UE), and reaching performance during no-stimulation trials separately for extension and flexion. For each participant and direction, no-stimulation data were pooled across the seven stimulation-target blocks and averaged for each metric. Metric values were log-transformed to improve residual normality and stabilize variance. A separate linear mixed-effects model was fitted for each metric, with FM-UE score, movement direction, and their interaction as fixed effects and participant as a random intercept. Associations with FM-UE were estimated for extension and flexion. Benjamini-Hochberg false discovery rate (FDR) correction was applied across the four metrics for the FM-UE × movement direction interactions and separately within each direction for the direction-specific associations [23]. Model estimates were converted from the log scale and reported as the percentage change in each metric per 10-point increase in FM-UE.

#### 2.5.3 fMRI preprocessing

Resting-state data were preprocessed using CONN toolbox 22.a and Statistical Parametric Mapping software (SPM12) [24,25]. Images were realigned and unwarped to correct for motion artifacts. Scans with framewise displacement greater than 0.9 mm or global blood-oxygen-level-dependent (BOLD) signal changes greater than five standard deviations were marked as outliers. Anatomical and BOLD data were normalized to MNI152 space using unified segmentation. The BOLD data were then resampled to 2-mm isotropic resolution and spatially smoothed using a Gaussian kernel with a 5-mm full width at half maximum (FWHM).

Scans were then denoised by regressing out five CompCor components each from white matter and cerebrospinal fluid, the six motion parameters and their first-order derivatives, outlier scans, session effects and their derivatives, and linear trends [26,27]. The residual time series were bandpass filtered between 0.008 and 0.09 Hz.

Before the resting-state group analyses, data were aligned by lesion side. Data from participants with left-hemisphere strokes (n = 7/19) were flipped across the midsagittal plane so that the ipsilesional hemisphere was represented on the right. Data from the two left-handed controls were also flipped so that the hemisphere ipsilateral to the dominant arm was represented on the right.

Task-based fMRI data were preprocessed using SPM12. Preprocessing included realignment, coregistration with the anatomical images, spatial smoothing with a 4-mm FWHM Gaussian kernel, and high-pass filtering at 0.01 Hz. T1-weighted images were segmented using FreeSurfer to support anatomical localization and registration of the functional data to each participant’s T1-and T2-weighted images [28]. Participant-level activation maps were then normalized to MNI152 space.

Data from participants who performed the task with the left arm were flipped across the midsagittal plane so that the hemisphere ipsilateral to the tested arm was represented on the right, providing consistent hemispheric alignment between the stroke and control groups.

#### 2.5.4 Regions of interest

Regions of interest (ROIs) were selected from cortical areas involved in upper-limb motor planning and movement. Bilateral PMd, PMv, and PPC corresponded to the regions targeted during TMS. Bilateral M1 was included as an additional region, with the ROIs centered on MNI coordinates corresponding to the hand-knob area (right: x = 35.85, y = −22.06, z = 55.84; left: x = −35.88, y = −23.94, z = 55.84). Each ROI was defined as a sphere with a 6.5-mm radius, with centers positioned slightly beneath the cortical surface to maintain gray-matter coverage and reduce overlap between adjacent regions.

#### 2.5.5 Task-based fMRI and group comparisons

Task-based fMRI data were analyzed separately for motor planning and movement. A general linear model (GLM) was constructed for each participant in SPM12, with separate regressors for the 3-s directional cue and the subsequent 3-s joystick movement. Leftward and rightward trials were combined within each regressor. Since the joystick task was performed primarily through wrist movement, leftward and rightward movements did not correspond directly to the extension and flexion movements performed during the KINARM task but were intended to be as feasible as possible in the MRI environment for people with motor impairment, and to simply model the motor planning aspect of reaching to one of two potential targets.

Separate contrast maps were generated for planning and movement relative to fixation. Individual maps were then thresholded using a whole-brain FDR-corrected criterion of *p_FDR_* < 0.05. The thresholded maps were then averaged within the stroke and control groups and projected onto the MNI152 cortical surface using AFNI/SUMA [29,30]. The resulting maps showed the spatial distribution of task-related activity within each group.

Group differences were tested within the predefined ROIs. For each participant, the peak *t* value was extracted from the unthresholded contrast map within each ROI separately for planning and movement. Peak *t* values were compared between the stroke and control groups using two-tailed independent-samples *t* tests. The eight ROI comparisons within each phase were FDR-corrected, with significance defined as *p_FDR_* < 0.05.

#### 2.5.6 Resting-state functional connectivity analysis

Resting-state fMRI was used to compare connectivity across the predefined ROIs between the stroke and control groups.

For each participant, the mean denoised BOLD time series was extracted from each ROI. Fisher z-transformed semipartial correlations were calculated in both directions for every ROI pair. These correlations represented the unique relationship between two ROIs after accounting for the time series from the remaining regions. Because semipartial correlations are asymmetric, each ROI pair yielded two values, one for each direction.

At the group level, a second-level GLM compared ROI-to-ROI connectivity between the stroke and control groups. A separate analysis within the stroke group used one-sample *t* tests to identify connections with mean connectivity significantly above or below zero.

Broader connectivity patterns were identified using CONN’s parametric functional network connectivity approach [31]. Complete-linkage hierarchical clustering grouped the ROIs based on anatomical proximity and similarity in their connectivity with the other regions. Multivariate *F* tests evaluated connectivity within and between the resulting ROI clusters. Clusters were considered significant at cluster-level *p_FDR_* < 0.05. Within each significant cluster, directional connections meeting a connection-level threshold of *p* < 0.05 were used to characterize the connectivity pattern. The same criteria were applied to the between-group and within-stroke analyses.

#### 2.5.7 Analysis of factors associated with TMS effects

To better understand the clinical and neural correlates of TMS effects on movement, we explored associations of the stimulation response with motor impairment and the resting-state connectivity patterns identified within the stroke group. Because TMS was delivered near the transition from planning to movement, we also tested its relationship with the change in task-evoked activity at the stimulated region across these phases using task-based fMRI.

TMS responses were calculated from the primary KINARM analysis using TDE and absolute LDJ as measures of trajectory straightness and smoothness, respectively. For each participant, movement direction, stimulation target, and intensity, the percentage change from PCS during no stimulation was subtracted from the corresponding change during subthreshold or suprathreshold stimulation. Within each movement direction, TDE and LDJ responses were separately converted to z scores and then averaged. Higher values of this movement-quality response score indicated poorer movement quality with stimulation.

Separate linear mixed-effects models were fitted for extension and flexion using maximum-likelihood estimation. The base model included stimulation target and intensity, categorized as subthreshold or suprathreshold, as fixed effects, and participant as a random intercept to account for repeated observations.

Motor impairment was represented by FM-UE score. Resting-state connectivity was summarized using one score for each significant cluster identified within the stroke group. Connections showing negative connectivity were sign-reversed so that higher values consistently represented stronger expression of the cluster’s connectivity pattern. Each directional semipartial correlation was standardized across participants, and the standardized values were averaged within each cluster. The resulting cluster score was standardized again across participants. FM-UE and each resting-state cluster score were evaluated in separate models.

The change in task-evoked activity was calculated for each participant and ROI by subtracting the planning phase peak t value from the movement phase peak t value. Each stroke participant’s value was standardized using the control group mean and standard deviation for the same ROI. Scores for bilateral PMd, PMv, and PPC were matched to stimulation of the same region and hemisphere. The control-referenced score and its interaction with stimulation intensity were added to the base model together.

Each clinical or imaging measure was evaluated by comparing the model including that measure with the base model. Model comparisons used the likelihood-ratio (*LR*) statistic, and significance was assessed using 10,000 participant-level permutations. During permutation, all predictor values belonging to a participant were reassigned together, preserving the repeated observations across stimulation targets and intensities.

## 3. Results

### 3.1 Direction-dependent effects of TMS on reaching

We assessed the effects of TMS on reach kinematics using measures of trajectory straightness (PLR and TDE) and smoothness (LDJ and SAL). Higher values indicated poorer movement quality for all four metrics. The mean RMT for the unaffected arm was 77.9 ± 19.4% of maximum stimulator output (range = 45–100%). For six participants, RMT exceeded 83% of maximum stimulator output, and suprathreshold stimulation was capped at 100% output. TMS began 100 ms before each participant’s calculated reaction time (mean = 369.3 ± 84.8 ms; range = 227–529 ms). Previous work supported PCS as a control target because it lay outside the principal task-evoked activation pattern and showed no systematic kinematic differences from the other targets during no stimulation [15].

The extension reach and group mean trajectories for each stimulation target and condition are shown in Figures 3a and 3b, respectively. Differences from PCS were most evident during suprathreshold stimulation, particularly at PMd and PPC, where the trajectories generally followed a more direct path.

**Figure 3.**
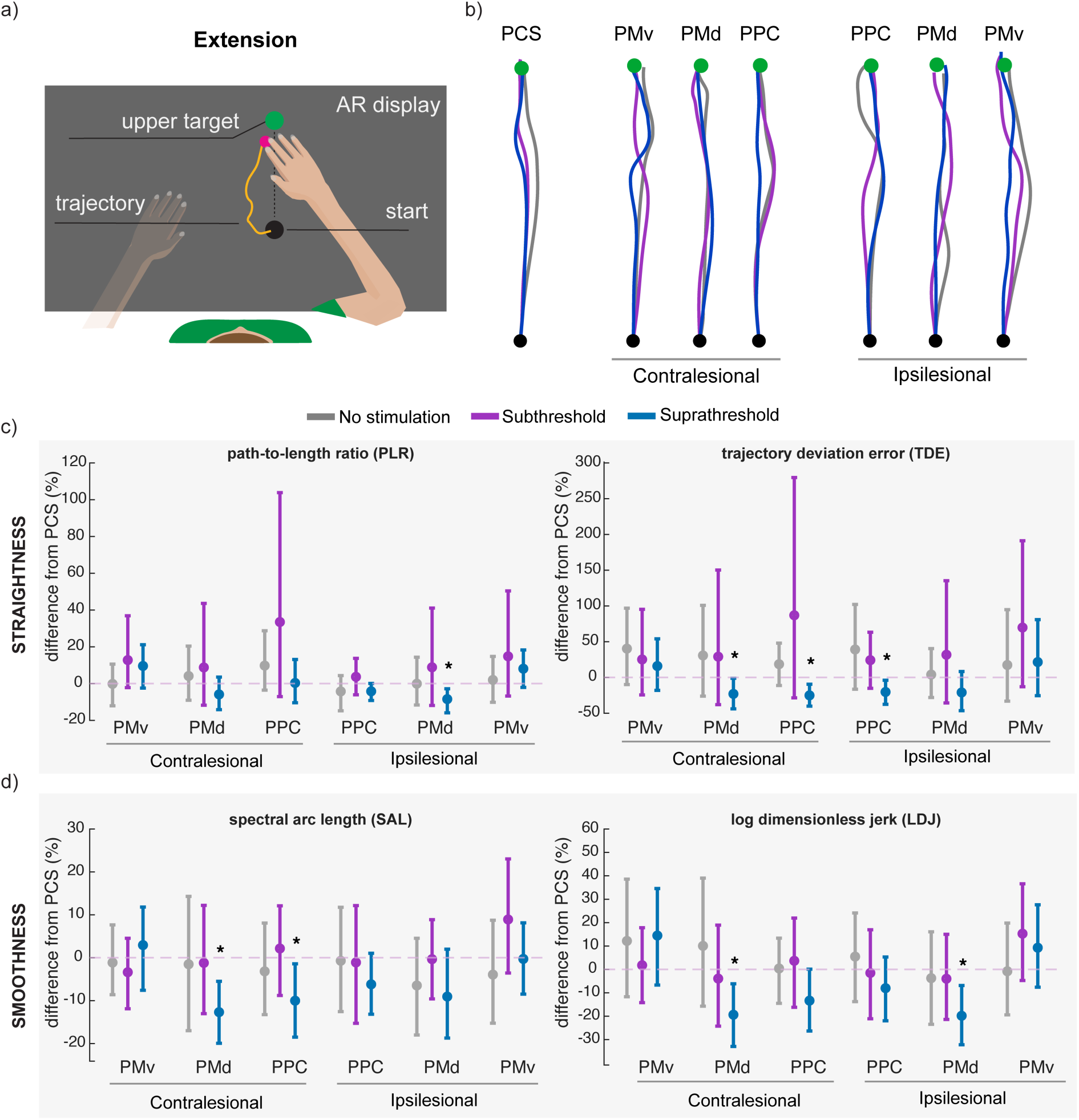
Effects of TMS on extension reaches. (a) Extension reach from the central starting position toward the upper, distal target. (b) Group-mean extension trajectories at each stimulation target during no stimulation (gray), subthreshold stimulation (purple), and suprathreshold stimulation (blue). Black and green circles indicate the starting position and target, respectively. (c) Percentage changes relative to PCS for the trajectory-straightness measures PLR and TDE. (d) Percentage changes relative to PCS for the smoothness measures SAL and LDJ. Asterisks indicate pointwise 95% bootstrap CIs excluding zero. Abbreviations: PMv, ventral premotor cortex; PMd, dorsal premotor cortex; PPC, posterior parietal cortex; PCS, postcentral sulcus.

Quantitative effects on trajectory straightness and smoothness are shown in Figures 3c and 3d, respectively. Extension-related changes were confined to suprathreshold stimulation and involved PMd and PPC in both hemispheres. In the contralesional hemisphere, which was ipsilateral to the affected arm, PMd stimulation reduced TDE by 23.0%, SAL by 12.7%, and LDJ by 19.4%, demonstrating improvements in both straightness and smoothness relative to PCS. PPC stimulation produced a similar effect, reducing TDE by 25.1% and SAL by 10.0%.

In the ipsilesional hemisphere, PPC stimulation reduced TDE by 20.3%. PMd stimulation reduced PLR by 8.5% and LDJ by 19.8%, corresponding to more efficient (shorter) and smoother trajectories. The 95% CIs for all reported effects excluded zero. No extension-related effects were observed at PMv in either hemisphere or during subthreshold stimulation. During extension, none of the stimulation targets differed from PCS in the no-stimulation condition. All 95% bootstrap CIs included zero.

The flexion reach and group mean trajectories for each stimulation target and condition are shown in Figures 4a and 4b, respectively. The no-stimulation trajectories were already relatively straight, and the trajectories remained largely overlapping across stimulation conditions. Thus, flexion performance showed little visual evidence of stimulation-related change.

**Figure 4.**
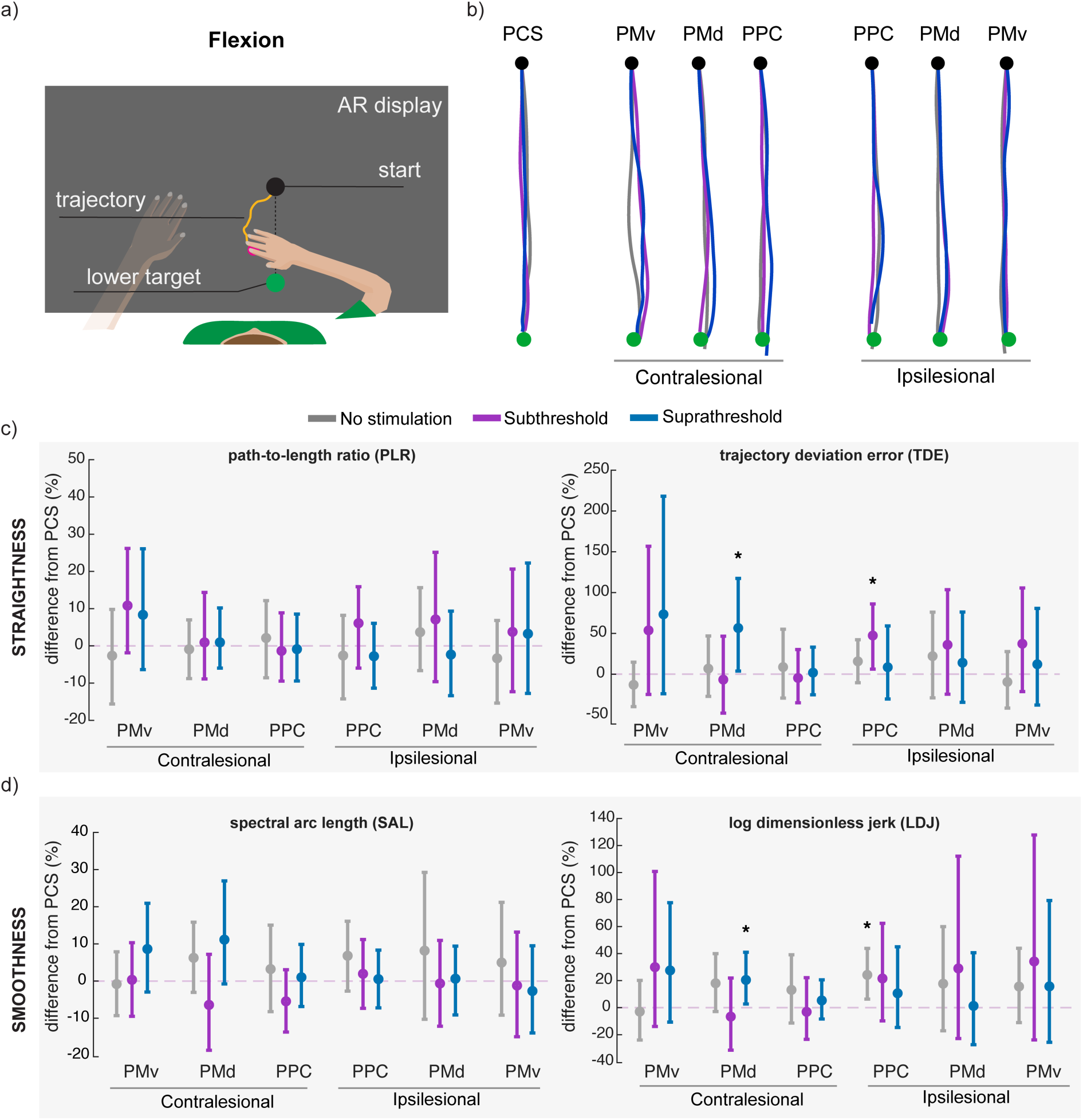
Effects of TMS on flexion reaches. (a) Flexion reach from the central starting position toward the lower, proximal target. (b) Group-mean flexion trajectories at each stimulation target during no stimulation (gray), subthreshold stimulation (purple), and suprathreshold stimulation (blue). Black and green circles indicate the starting position and target, respectively. (c) Percentage changes relative to PCS for the trajectory straightness measures PLR and TDE. (d) Percentage changes relative to PCS for the smoothness measures SAL and LDJ. Asterisks indicate pointwise 95% bootstrap CIs excluding zero. Abbreviations: PMv, ventral premotor cortex; PMd, dorsal premotor cortex; PPC, posterior parietal cortex; PCS, postcentral sulcus.

Effects on trajectory straightness and smoothness are shown in Figures 4c and 4d, respectively. Fewer differences from PCS were observed during flexion than during extension. In the contralesional hemisphere, suprathreshold PMd stimulation increased TDE by 56.5% and LDJ by 21.3%, showing greater trajectory deviation and reduced smoothness relative to PCS. No other contralesional differences from PCS were observed during flexion.

In the ipsilesional hemisphere, subthreshold PPC stimulation increased TDE by 47.4%, while no effects were observed at PMd or PMv. In the no-stimulation condition, LDJ was 24.0% higher at ipsilesional PPC than at PCS, while PLR, TDE, and SAL did not differ. This finding was interpreted cautiously because block order or fatigue may have contributed to the difference. The 95% CIs for all reported effects excluded zero.

To directly test the effects of stimulation by movement direction, we compared the changes relative to PCS stimulation between extension and flexion within participants. The clearest direction-dependent effect was observed at contralesional PMd during suprathreshold stimulation. Compared with flexion, extension showed more favorable PCS-normalized changes for PLR (mean difference = −13.8 percentage points), TDE (−101.4 percentage points), SAL (−20.9 percentage points), and LDJ (−47.2 percentage points). The 95% bootstrap CIs for all four differences excluded zero. This pattern was consistent with the separate direction analyses: suprathreshold contralesional PMd stimulation reduced extension TDE, SAL, and LDJ and increased flexion TDE and LDJ relative to PCS.

Suprathreshold contralesional PPC stimulation also improved extension performance, reducing TDE and SAL relative to PCS. However, the 95% bootstrap CIs for the direct comparisons all included zero. No other suprathreshold target showed a direct extension–flexion difference. Additional direction differences were observed during subthreshold stimulation for TDE at ipsilesional PPC and SAL at contralesional PPC, but each was limited to a single metric.

### 3.2 Baseline reaching performance and motor function

Baseline reaching performance was associated with motor impairment severity, as measured by the FM-UE (Figure S1). Associations with FM-UE were significantly different between extension and flexion for all four metrics (*p_FDR_* ≤ 0.029) and were stronger during extension. During extension, a 10-point higher FM-UE score was associated with 15.0% lower PLR (*p_FDR_* < 0.001), 28.5% lower TDE (*p_FDR_* < 0.001), and 12.3% lower absolute LDJ (*p_FDR_* = 0.007). Absolute SAL showed opposite-signed associations with FM-UE during extension and flexion, although neither direction-specific association was significant. During flexion, a 10-point higher FM-UE score was associated with 16.1% lower TDE (*p_FDR_* = 0.048). No significant associations were found for flexion PLR or LDJ.

### 3.3 Task-evoked activity in stroke and control groups

A broad network of motor and multisensory regions was active during the joystick task in both groups (Figure 5). During the planning phase, activity was observed in motor cortex and PMd in the hemisphere contralateral to the tested arm (ipsilesional in the stroke group), as well as in the supplementary motor area, bilateral PMv and PPC, and visual cortex (Figure 5a).

**Figure 5.**
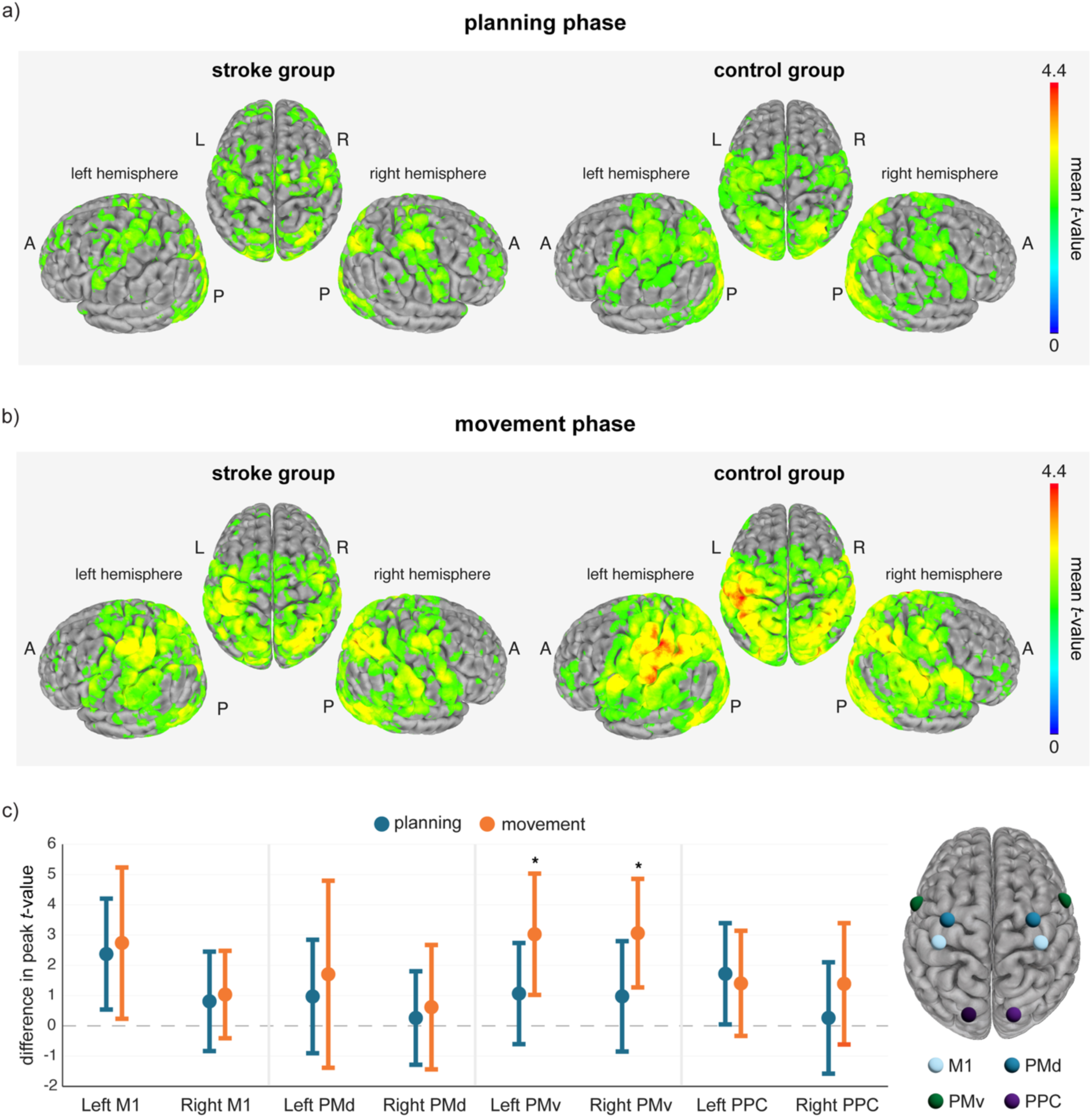
Task-based fMRI activation in the stroke and control groups. Group mean activation maps are shown during (a) planning and (b) joystick movement using the same scale. (c) Differences in peak t values between the groups (control − stroke) within each ROI. Positive values indicate higher activity in controls. Error bars show 95% CIs, and asterisks indicate *p_FDR_* < 0.05. The brain schematic shows the ROI locations. Throughout the figure, left and right refer to the aligned display, with the hemisphere contralateral to the tested arm shown on the left and the hemisphere ipsilateral to the tested arm shown on the right. Abbreviations: M1, primary motor cortex; PMv, ventral premotor cortex; PMd, dorsal premotor cortex; PPC, posterior parietal cortex; A, anterior; P, posterior.

During the movement phase, activity was more extensive across the sensorimotor cortex contralateral to the tested arm and bilateral premotor and parietal regions (Figure 5b). Bilateral cerebellar activity was also present during both phases. Participants with stroke showed a broadly similar spatial pattern, although mean *t* values appeared lower, particularly during movement. Within the defined ROIs, no group differences were found during planning after FDR correction.

During the movement phase, controls showed higher peak activation in PMv contralateral to the tested arm (control–stroke difference = 3.03, *t*(33) = 3.08, *p_FDR_* = 0.017) and PMv ipsilateral to the tested arm (difference = 3.06, *t*(33) = 3.47, *p_FDR_* = 0.012).

### 3.4 Functional connectivity patterns

Resting-state functional connectivity was compared between the stroke and control groups across ROIs. Figure 6a shows the direction and strength of each group difference, with *p_FDR_* values shown within the cells. After FDR correction, only interhemispheric PPC connectivity differed between groups and was stronger in the stroke group, *F*(1, 42) = 10.39, *p_FDR_* = 0.032. This difference was significant from ipsilesional to contralesional PPC, *t*(42) = 3.24, *p_FDR_* = 0.017, and from contralesional to ipsilesional PPC, *t*(42) = 3.17, *p_FDR_* = 0.020. No other connections differed significantly between groups.

**Figure 6.**
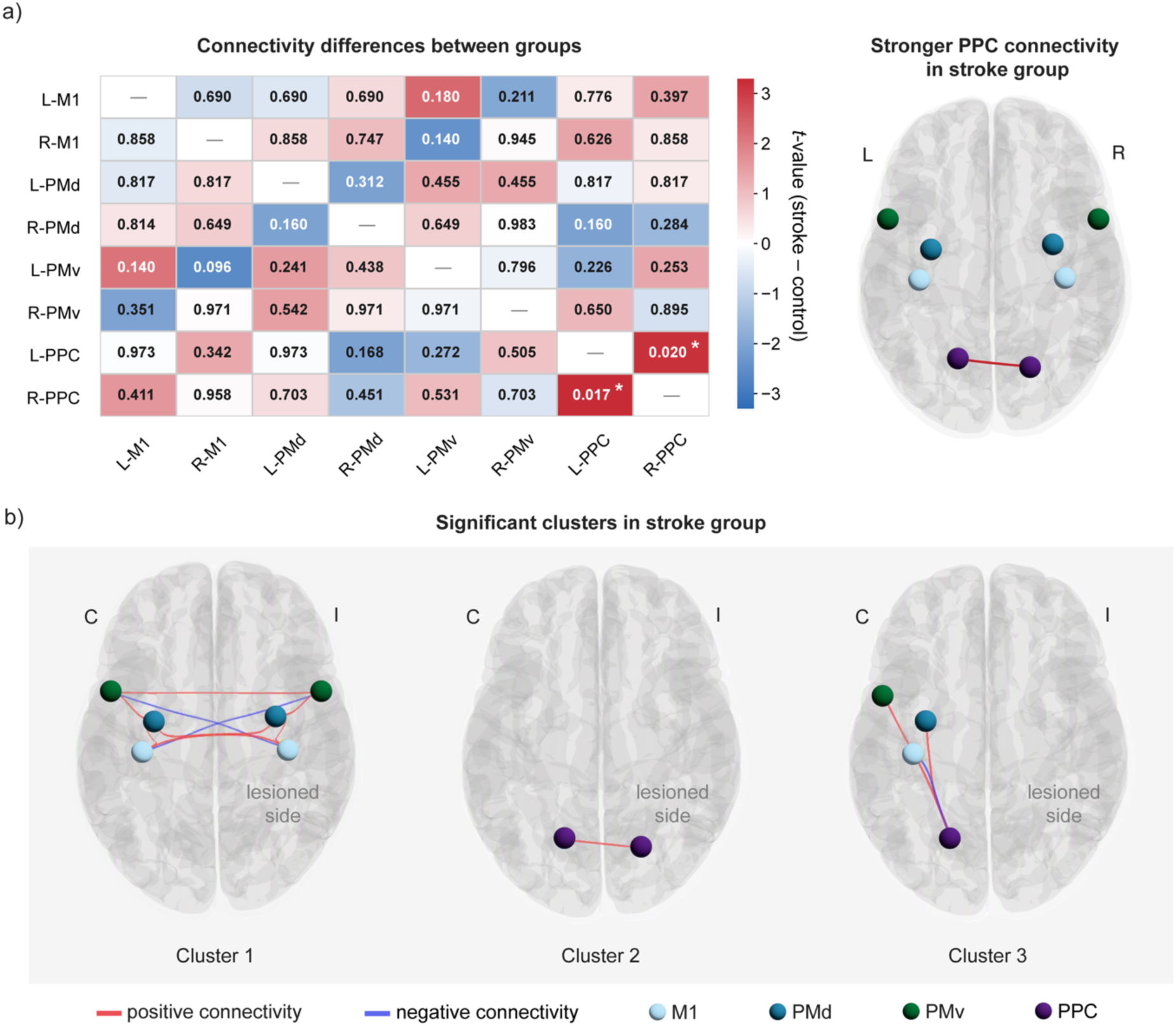
Resting-state functional connectivity. (a) Connectivity differences between the stroke and control groups across the ROIs. Heatmap color represents the t value for the stroke–control comparison, with red indicating stronger connectivity in the stroke group and blue indicating stronger connectivity in controls. Cell values show connection-level *p_FDR_* values. Asterisks indicate *p_FDR_* < 0.05. In the aligned stroke data, L and R correspond to the contralesional and ipsilesional hemispheres, respectively. The brain diagram shows the significantly stronger interhemispheric PPC connectivity in the stroke group in both directions. (b) Connections contributing to the significant clusters identified within the stroke group. Red and blue lines indicate positive and negative connectivity, respectively. Abbreviations: C, contralesional hemisphere; I, ipsilesional hemisphere; M1, primary motor cortex; PMv, ventral premotor cortex; PMd, dorsal premotor cortex; PPC, posterior parietal cortex.

Three significant connectivity clusters were identified within the stroke group (Figure 6b, Table 2). Cluster 1 included M1, PMd, and PMv in both hemispheres, *F*(2, 17) = 203.61, *p_FDR_* < 0.001. Positive connectivity was found between homologous regions across hemispheres, including bilateral M1, PMd, and PMv, and among motor and premotor regions within each hemisphere. The cluster also included negative connectivity between M1 and PMv across hemispheres.

**Table 2.** Resting-state functional connectivity clusters in the stroke group.

| Cluster | Connections | Directional<br>$t(18)$ | Connection $p_{FDR}$ | Cluster<br>statistic | Cluster<br>$p_{FDR}$ |
| --- | --- | --- | --- | --- | --- |
| 1 | cPMd ↔ iPMd | 8.34 / 7.89 | < 0.001 / < 0.001 |  |  |
|  | cM1 ↔ iM1 | 7.29 / 7.37 | < 0.001 / < 0.001 |  |  |
|  | cM1 ↔ cPMd | 6.30 / 6.28 | < 0.001 / < 0.001 |  |  |
|  | cPMv ↔ iPMv | 5.91 / 6.30 | < 0.001 / < 0.001 |  |  |
| | iM1 ↔ iPMd | 5.42 / 5.58 | < 0.001 / < 0.001 | $F(2,17) = 203.61$ | < 0.001 |
|  | iPMd ↔ iPMv | 4.46 / 4.66 | < 0.001 / < 0.001 |  |  |
|  | cPMd ↔ cPMv | 4.23 / 4.19 | 0.001 / 0.002 |  |  |
|  | iM1 ↔ cPMv | -3.29 / -2.89 | 0.009 / 0.019 |  |  |
|  | cM1 ↔ iPMv | -2.41 / -2.37 | 0.063 / 0.068 |  |  |
| 2 | cPPC ↔ iPPC | 9.33 / 9.30 | < 0.001 / < 0.001 | $F(1,18) = 88.28$ | < 0.001 |
| 3 | cPMv ↔ cPPC | 2.84 / 2.93 | 0.019 / 0.031 |  |  |
| | cPMd ↔ cPPC | 2.74 / 2.71 | 0.023 / 0.033 | $F(2,17) = 4.10$ | 0.035 |
|  | cPPC → cM1 | -2.23 | 0.067 |  |  |

For bidirectional connections, statistics are reported in the order shown (first region → second region / second region → first region). c, contralesional; i, ipsilesional.

Cluster 2 consisted of positive connectivity between the contralesional and ipsilesional PPC, *F*(1, 18) = 88.28, *p_FDR_* < 0.001. Both directional connections contributed to the cluster.

Cluster 3 included connections in the contralesional hemisphere between PPC, PMd, PMv, and M1, *F*(2, 17) = 4.10, *p_FDR_* = 0.035. The cluster was characterized by positive bidirectional PPC connectivity with PMd and PMv. Directional statistics for all connections contributing to the three clusters are reported in Table 2.

### 3.5 Clinical and neural correlates of TMS effects

We aimed to explain the TMS response based on the severity of motor impairment and the neural patterns measured with resting-state and task-based fMRI. Higher FM-UE scores were associated with better reaching performance without stimulation but not with the stimulation-specific response during extension (*LR* = 2.11, *p* = 0.204) or flexion (*LR* = 0.93, *p* = 0.384).

The resting-state cluster scores were also unrelated to TMS response. During extension, no associations were found for the motor–premotor connections in cluster 1 (*LR* = 0.93, *p* = 0.413), cluster 2 (*LR* = 0.40, *p* = 0.601), or cluster 3 (*LR* = 2.46, *p* = 0.179). The same was found during flexion for cluster 1 (*LR* = 1.01, *p* = 0.376), the interhemispheric PPC connection in cluster 2 (*LR* = 0.82, *p* = 0.424), and the contralesional frontoparietal connections in cluster 3 (*LR* = 1.38, *p* = 0.301).

The planning-to-movement change in task-evoked activity showed a different pattern. During extension, jointly adding the control-referenced activity-change score and its interaction with stimulation intensity improved model fit (*LR* = 9.75, participant-level permutation *p* = 0.036; Figure 7a). The fitted associations were positive during both subthreshold (*β* = 0.18) and suprathreshold stimulation (*β* = 0.31), with no evidence that the slopes differed by intensity (interaction *β* = 0.13, *p* = 0.383). In this exploratory analysis, lower control-referenced planning-to-movement activity changes were associated with more favorable responses to TMS during extension movements.

**Figure 7.**
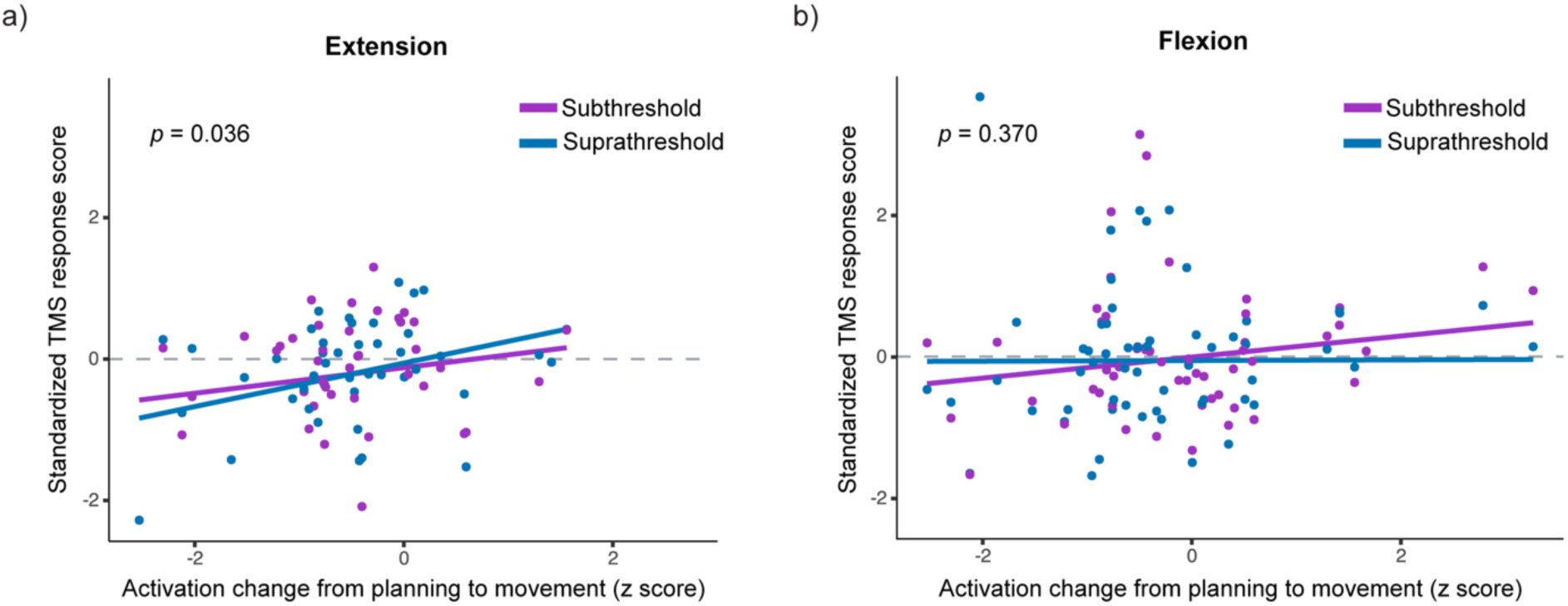
Association between task-related activation change and TMS response. Activation change from planning to movement at each stimulated ROI was standardized using the control-group distribution for the corresponding region and plotted against the TMS response during (a) extension and (b) flexion. Each point represents one participant, stimulation site, and intensity observation. Purple and blue indicate subthreshold and suprathreshold stimulation, respectively, and lines show the fitted associations from the mixed-effects model. Higher TMS response scores indicate poorer movement quality with stimulation. The displayed *p* values are participant-level permutation values from the joint likelihood-ratio test of the activity-change score and its interaction with intensity.

No corresponding relationship was found during flexion (*LR* = 2.12, participant-level permutation *p* = 0.370; Figure 7b).

## 4. Discussion

The goal of this study was to understand how cortical regions involved in planning and guiding a reach contribute to affected arm movement after stroke, and how their contributions change with the demands of the movement being planned. We combined TMS delivered before movement with a planar reaching task to probe these contributions bilaterally in premotor cortex (PMd and PMv) and the dorsomedial SPL within PPC, regions with established roles in reach planning and guidance [32–34]. To place the stimulation effects in the context of cortical function after stroke, we used task-based fMRI to map task-evoked activity during planning and movement and quantify planning-to-movement activity changes across cortical regions. Resting-state fMRI was used to characterize connectivity patterns among premotor, parietal, and primary motor regions.

Although all movements were multijoint reaches performed within the same planar KINARM task, target location changed both reach direction and the required pattern of shoulder and elbow motion. Performance during reaches to the distal target, which primarily required elbow extension, was more strongly related to motor impairment. Suprathreshold stimulation of PMd and PPC in both hemispheres improved trajectory straightness or smoothness during these reaches. At contralesional PMd, the same stimulation had opposite effects across reach directions, with favorable effects for distal reaches and unfavorable effects for proximal reaches, which primarily required elbow flexion. Compared with controls, interhemispheric PPC connectivity was stronger in the stroke group. Within the stroke group, significant resting-state clusters were either bilateral or confined to the contralesional hemisphere; none was confined to the ipsilesional hemisphere. In contrast, the previous control cohort showed within-hemisphere connectivity in both hemispheres [15]. In the exploratory analyses, smaller planning-to-movement activity increases relative to controls were associated with more favorable kinematic effects of TMS during distal reaches.

The stronger relationship between performance and impairment during distal reaches suggests that these movements rely more heavily on residual motor capacity. Similar directional differences have been reported after stroke. Outward movements that include elbow extension are often slower and more curved. These features relate to different aspects of motor impairment, with movement speed associated with force-generating capacity and path curvature associated with the ability to control the shoulder and elbow independently [35]. Even when the arm is supported, outward movements show greater disruption of multijoint torque coordination than inward movements, and the relationship between this coordination and FM-UE is stronger for outward movements [36].

Within the premotor cortices (Brodmann area 6), changes in reaching were observed after PMd stimulation but not after PMv stimulation. At ipsilesional PMd, pointwise estimates indicated shorter and smoother trajectories toward the distal target, although direct comparisons did not support a difference between reach directions. The opposite effects of contralesional PMd stimulation across reach directions may partly reflect baseline performance. Trajectories toward the proximal target were already relatively straight, leaving limited room for further improvement. Perturbing PMd during the preparation of these movements may instead have disrupted a relatively preserved control strategy. Distal reaches, by contrast, may have offered greater scope for stimulation to improve trajectory straightness and smoothness.

The involvement of PMd in both hemispheres aligns with evidence that the integrity of its descending fibers is specifically related to motor behaviors requiring interjoint coordination and visuomotor integration after stroke [37]. Unger et al. also found that stimulation of ipsilesional premotor cortex increased connectivity between the two premotor cortices in participants with moderate-to-severe stroke, and that this change was associated with recovery of proximal arm function [38]. Direct stimulation studies of contralesional PMd have likewise yielded different behavioral results. Lin et al. reported better reaching after contralesional PMd stimulation in participants with greater impairment [39]. Mustin et al. found no consistent group-level effect of contralesional PMd stimulation, although the effects of contralesional stimulation more broadly varied with the remaining structural integrity of the ipsilesional motor network [40]. Neither study compared different reach directions within the same task. In our study, the same contralesional PMd stimulation had opposite effects within participants depending on the reach being planned. This pattern was also different from our previous study of neurologically intact adults, in which PMd stimulation generally reduced reach efficiency and smoothness [15].

PMv stimulation did not alter reaching in the present study, despite affecting performance in the previous control cohort [15]. Casarotto et al. found that modulation of the PMv-to-M1 pathway changed the temporal coordination of precision grasping but not power grasping [41], while Turrini et al. found sustained changes in corticospinal excitability when PMv-to-M1 stimulation was paired with movement execution but not with movement planning [42]. Our task required neither grasp nor object manipulation, and stimulation began near the transition from planning to movement. The absence of a PMv effect may therefore be specific to the behavior and timing here.

PPC stimulation in either hemisphere also altered reaching, but only for particular combinations of movement direction and intensity. During distal reaches, suprathreshold contralesional PPC stimulation reduced deviation and increased smoothness, whereas suprathreshold ipsilesional stimulation reduced deviation. During proximal reaches, subthreshold ipsilesional stimulation instead increased deviation. The favorable suprathreshold effects contrast with our neurologically intact cohort, in which stimulation of the same target increased deviation and reduced smoothness [15]. Our target was located in the dorsomedial SPL, within a parietofrontal network involved in transporting the arm toward a spatial goal. Vesia et al. found that three-pulse, 10-Hz stimulation during reach planning altered remembered reach endpoints across several PPC regions. At the medial-posterior site, these effects were less restricted to the hand contralateral to the stimulated hemisphere [43], providing a plausible basis for the bilateral effects observed here. A separate study showed that functional interactions between medial PPC and M1 increased when the planned action required arm transport [44]. After stroke, TMS over contralesional SPL disrupted recovered hand sequencing [14], whereas stimulation of contralesional anterior intraparietal sulcus (aIPS) improved motor performance early after stroke [5] but impaired grasp accuracy in chronic stroke when interhemispheric aIPS connectivity was stronger [45]. Our findings extend this evidence to dorsomedial PPC during paretic-arm reaching and suggest that its contribution may vary with the movement being planned and stimulation intensity.

During both planning and movement, participants with stroke showed a spatial pattern of task-evoked activity similar to controls, but bilateral PMv activity was significantly lower during movement. Previous studies have reported reduced motor-network activity after stroke, greater PMv activity in well-recovered patients, and increased PMv activity after arm training [46–48]. These findings suggest that PMv activity varies with the task and stage of recovery and does not change uniformly after stroke.

At rest, the stronger interhemispheric PPC connectivity in the stroke group was consistent with altered interhemispheric PPC connectivity reported in chronic stroke, although the direction of these changes was not consistent across parietal regions [49]. Within the stroke group, the significant connections formed three broader patterns: bilateral motor-premotor, interhemispheric PPC, and contralesional parietofrontal connectivity. These within-group patterns are consistent with bilateral and contralesional connectivity reported in other studies. Zhang et al. found increased motor-network connectivity in the contralesional SPL compared with controls, and participants with stronger connectivity had higher FM-UE scores [50]. Lee et al. found that interhemispheric motor-network connectivity was disrupted early after stroke and was related to both residual motor function and subsequent improvement, with aIPS among the regions involved [51]. Among patients with extensive corticospinal tract damage, however, better performance has been related to stronger ipsilesional frontoparietal connectivity [52]. The relative importance of these pathways may therefore depend on the stage of recovery and remaining corticospinal integrity.

Among the clinical and imaging measures, the planning-to-movement change in task-evoked activity at the stimulated region was the only measure significantly associated with TMS response, and this association was limited to distal reaches. Smaller increases relative to controls were associated with more favorable responses, whereas increases that approached or exceeded the control pattern were associated with less favorable responses. This does not imply that lower activity is inherently beneficial. It suggests that the behavioral effect of TMS may depend partly on how activity in the stimulated region changes as movement begins. Bestmann et al. found that, during paretic-hand grip, contralesional PMd stimulation had a stronger facilitatory effect on posterior ipsilesional sensorimotor activity in participants with greater impairment [53]. PPC stimulation has likewise produced stronger neural and behavioral effects during an active motor task than at rest [54]. Resting-state connectivity was not associated with TMS response here, although previous work suggests that its relevance may vary by stimulation target [6]. Task-evoked activity at the targeted region may therefore contribute to variability in the response to TMS.

Several limitations should be acknowledged. Not every participant completed all study components, so analyses of associations between behavioral and imaging measures included fewer participants (n = 15 for resting-state fMRI and n = 13 for task-based fMRI). The timing of stimulation also relied on reaction times measured during practice and may not have remained equally aligned with movement onset if fatigue or practice changed performance over the session. Stimulation intensity was also based on RMT measured from M1 contralateral to the unaffected arm, and suprathreshold output was capped in six participants, potentially introducing variation in effective intensity. While use of RMT to scale stimulation strength is a standard practice, it may not account for different brain geometry and other variations that affect the effectiveness of TMS in regions beside the motor cortex. In addition, the KINARM task was limited to two planar reaching directions with the arm supported and did not capture the full complexity of natural reaching. However, premotor TMS has also altered trajectories during unconstrained reaching [55], supporting the relevance of our findings to less constrained movements. The fMRI association also requires caution. The joystick task relied primarily on wrist movement, and the KINARM task required planar arm reaches. The tasks were also completed on separate days. Despite these differences, both distinguished planning from execution, which was the feature central to the association analysis. Even so, fMRI does not measure cortical activity at the moment of stimulation.

Future studies could adjust pulse timing from trial to trial and compare repeated or alternative stimulation protocols. This would establish whether the immediate direction-dependent effects observed here can lead to lasting motor benefits. The long-term goal is to develop patient-specific stimulation protocols that can be combined with practice to improve voluntary motor function.

## 5. Conclusions

The contribution of premotor and parietal cortex to affected arm reaching varied by reach direction after stroke. The influence of bilateral PMd and dorsomedial PPC was most evident during reaches that drew more strongly on residual motor capacity. At contralesional PMd, the same perturbation had favorable effects during distal reaches and unfavorable effects during proximal reaches. The imaging results provided a broader cortical context. Compared with controls, the stroke group showed stronger interhemispheric PPC connectivity. The association with task-evoked activity further suggests that the response to stimulation may be related to how activity at the targeted region changes from planning to movement. These findings argue against selecting a stimulation target without considering the movement it is intended to influence. Neuromodulation after stroke may therefore be more effective when cortical targets are selected according to the movement and the network supporting that movement.

## List of abbreviations

aIPS: anterior intraparietal sulcus
BOLD: blood-oxygen-level-dependent
CI: confidence interval
EDC: extensor digitorum communis
EMG: electromyography
FDR: false discovery rate
FM-UE: Fugl-Meyer Assessment of the Upper Extremity
fMRI: functional magnetic resonance imaging
FWHM: full width at half maximum
GLM: general linear model
LDJ: log-dimensionless jerk
LR: likelihood ratio
M1: primary motor cortex
MEP: motor-evoked potential
MNI: Montreal Neurological Institute
MRI: magnetic resonance imaging
PCS: postcentral sulcus
PLR: path-to-length ratio
PMd: dorsal premotor cortex
PMv: ventral premotor cortex
PPC: posterior parietal cortex
RMT: resting motor threshold
ROI: region of interest
rsfMRI: resting-state functional magnetic resonance imaging
SAL: spectral arc length
SD: standard deviation
SPL: superior parietal lobule
TDE: trajectory deviation error
tDCS: transcranial direct current stimulation
TE: echo time
TMS: transcranial magnetic stimulation
TR: repetition time

## Declarations

### Ethics approval and consent to participate

Study protocols were approved by the Institutional Review Boards of the University of Pittsburgh (study #19080097) and Veterans Affairs Pittsburgh Healthcare System (study #1617353). All participants provided written informed consent before enrollment.

### Consent for publication

All participants provided written informed consent for the publication of their deidentified data.

### Availability of data and materials

The data that support the findings of this study are available on request from the corresponding author. The data are not publicly available due to privacy or ethical restrictions.

### Competing interests

The authors declare that they have no competing interests.

### Funding

This work was supported by the Veterans Health Administration Rehabilitation Research & Development Program Merit Review Award (#I01 RX003511). The contents do not represent the views of the U.S. Department of Veterans Affairs or the United States Government.

## Authors’ contributions

**GH:** Conceptualization, Data curation, Formal analysis, Investigation, Methodology, Visualization, Writing – original draft, Writing – review & editing;

**RMF:** Conceptualization, Data curation, Formal analysis, Investigation, Methodology, Writing – review & editing; **JM:** Investigation, Methodology, Software, Writing – review & editing; **AB:** Investigation, Project administration, Resources, Writing – review & editing; **XF:** Investigation, Methodology, Software, Writing – review & editing; **JLC:** Resources, Supervision, Writing – review & editing; **GM:** Methodology, Writing – review & editing; **LZ:** Methodology, Supervision, Writing – review & editing; **FL:** Conceptualization, Data curation, Formal analysis, Investigation, Methodology, Software, Visualization, Writing – review & editing; **GFW:** Conceptualization, Funding acquisition, Methodology, Project administration, Resources, Supervision, Writing – review & editing. All authors read and approved the final manuscript.

## Acknowledgements

We would like to thank Tyler Madonna and Tyler Simpson for technical support with the KINARM robot and software. We also thank Debbie Harrington and Sydney Bader for their help with participant recruitment. We are grateful to Dr. Peter Strick for discussions regarding the targeting of the premotor region – PMv in particular – and to Dr. Mike Urbin for valuable discussions on the study design and interpretation.

## Notes

### Competing Interest Statement

The authors have declared no competing interest.

### Clinical Trial

NCT04286516

## References

1. Ahmed I, Mustafaoglu R, Rossi S, Cavdar FA, Agyenkwa SK, Pang MYC, et al. Non-invasive brain stimulation techniques for the improvement of upper limb motor function and performance in activities of daily living after stroke: a systematic review and network meta-analysis. Arch Phys Med Rehabil. 2023;104(10):1683–97.

2. Ovadia-Caro S, Khalil AA, Sehm B, Villringer A, Nikulin VV, Nazarova M. Predicting the response to non-invasive brain stimulation in stroke. Front Neurol. 2019;10:302.

3. Hildesheim FE, Silver AN, Dominguez-Vargas AU, Andrushko JW, Edwards JD, Dancause N, et al. Predicting individual treatment response to rTMS for motor recovery after stroke: a review and the CanStim perspective. Front Rehabil Sci. 2022;3:795335.

4. Koch G, Rothwell JC. TMS investigations into the task-dependent functional interplay between human posterior parietal and motor cortex. Behav Brain Res. 2009;202(2):147–52.

5. Tscherpel C, Hensel L, Lemberg K, Vollmer M, Volz LJ, Fink GR, et al. The differential roles of contralesional frontoparietal areas in cortical reorganization after stroke. Brain Stimul. 2020;13(3):614–24.

6. Hensel L, Bonkhoff AK, Paul T, Tscherpel C, Lange F, Viswanathan S, et al. The role of contralesional regions for post-stroke movements revealed by dynamic connectivity and TMS interference. NeuroImage Clin. 2025;47:103825.

7. Yao J, Drogos J, Veltink F, Anderson C, Concha Urday Zaa J, Hanson LI, et al. The effect of transcranial direct current stimulation on the expression of the flexor synergy in the paretic arm in chronic stroke is dependent on shoulder abduction loading. Front Hum Neurosci. 2015;9:262.

8. Gertz H, Lingnau A, Fiehler K. Decoding movement goals from the fronto-parietal reach network. Front Hum Neurosci. 2017;11:84.

9. Pilacinski A, Lindner A. Distinct contributions of human posterior parietal and dorsal premotor cortex to reach trajectory planning. Sci Rep. 2019;9(1):1962.

10. Davare M, Zénon A, Desmurget M, Olivier E. Dissociable contribution of the parietal and frontal cortex to coding movement direction and amplitude. Front Hum Neurosci. 2015;9:241.

11. Falaki A, Quessy S, Dancause N. Differential modulation of local field potentials in the primary and premotor cortices during ipsilateral and contralateral reach to grasp in macaque monkeys. J Neurosci. 2024;44(21):e1161232024.

12. Wiemer VM, Paul T, Hensel L, Cieslak M, Tscherpel C, Grefkes C, et al. Structural reserve-dependent cortical rerouting of motor control after stroke. Brain. 2026;149(7):2505–15.

13. Sankarasubramanian V, Machado AG, Conforto AB, Potter-Baker KA, Cunningham DA, Varnerin NM, et al. Inhibition versus facilitation of contralesional motor cortices in stroke: deriving a model to tailor brain stimulation. Clin Neurophysiol. 2017;128(6):892–902.

14. Lotze M, Markert J, Sauseng P, Hoppe J, Plewnia C, Gerloff C. The role of multiple contralesional motor areas for complex hand movements after internal capsular lesion. J Neurosci. 2006;26(22):6096–102.

15. Haddadshargh G, De Freitas RM, Mak J, Boos A, Fang X, Collinger JL, et al. Mapping the Causal Roles of Non-Primary Motor Areas in Human Reach Planning and Execution. Hum Brain Mapp. 2026 Feb 1;47(2):e70465. doi:10.1002/hbm.70465

16. Yarkoni T, Poldrack RA, Nichols TE, Van Essen DC, Wager TD. Large-scale automated synthesis of human functional neuroimaging data. Nat Methods. 2011;8(8):665–70.

17. Jenkinson M, Bannister P, Brady M, Smith S. Improved optimization for the robust and accurate linear registration and motion correction of brain images. Neuroimage. 2002;17(2):825–41.

18. Jenkinson M, Smith S. A global optimisation method for robust affine registration of brain images. Med Image Anal. 2001;5(2):143–56.

19. Awiszus F, Borckardt J. TMS motor threshold assessment tool (MTAT 2.0). Brain Stimul Lab Med Univ S C USA. 2011.

20. Balasubramanian S, Melendez-Calderon A, Burdet E. A robust and sensitive metric for quantifying movement smoothness. IEEE Trans Biomed Eng. 2011;59(8):2126– 36.

21. Powell MP, Verma N, Sorensen E, Carranza E, Boos A, Fields DP, et al. Epidural stimulation of the cervical spinal cord for post-stroke upper-limb paresis. Nat Med. 2023;29(3):689–99.

22. McAuley JH, Rothwell JC, Marsden CD. Frequency peaks of tremor, muscle vibration and electromyographic activity at 10 Hz, 20 Hz and 40 Hz during human finger muscle contraction may reflect rhythmicities of central neural firing. Exp Brain Res. 1997;114:525–41.

23. Benjamini Y, Hochberg Y. Controlling the False Discovery Rate: A Practical and Powerful Approach to Multiple Testing. J R Stat Soc Ser B Stat Methodol. 1995 Jan 1;57(1):289–300. doi:10.1111/j.2517-6161.1995.tb02031.x

24. Whitfield-Gabrieli S, Nieto-Castanon A. *conn*: a functional connectivity toolbox for correlated and anticorrelated brain networks. Brain Connect. 2012 Jun;2(3):125–41. doi:10.1089/brain.2012.0073

25. Friston KJ, Ashburner JT, Kiebel SJ, Nichols TE, Penny WD, editors. Statistical parametric mapping: the analysis of functional brain images. Amsterdam: Academic Press; 2007.

26. Behzadi Y, Restom K, Liau J, Liu TT. A component based noise correction method (CompCor) for BOLD and perfusion based fMRI. Neuroimage. 2007;37(1):90–101.

27. Power JD, Mitra A, Laumann TO, Snyder AZ, Schlaggar BL, Petersen SE. Methods to detect, characterize, and remove motion artifact in resting state fMRI. Neuroimage. 2014;84:320–41.

28. Fischl B. FreeSurfer. Neuroimage. 2012;62(2):774–81.

29. Cox RW. AFNI: software for analysis and visualization of functional magnetic resonance neuroimages. Comput Biomed Res. 1996;29(3):162–73.

30. Saad ZS, Reynolds RC. Suma. Neuroimage. 2012;62(2):768–73.

31. Nieto-Castanon A. Handbook of functional connectivity magnetic resonance imaging methods in CONN. Boston (MA): Hilbert Press; 2020. doi:10.56441/hilbertpress.2207.6598.

32. Hoshi E, Tanji J. Distinctions between dorsal and ventral premotor areas: anatomical connectivity and functional properties. Curr Opin Neurobiol. 2007;17(2):234–42.

33. Bernier PM, Cieslak M, Grafton ST. Effector selection precedes reach planning in the dorsal parietofrontal cortex. J Neurophysiol. 2012 Jul 1;108(1):57–68. doi:10.1152/jn.00011.2012

34. Desmurget M, Epstein CM, Turner RS, Prablanc C, Alexander GE, Grafton ST. Role of the posterior parietal cortex in updating reaching movements to a visual target. Nat Neurosci. 1999;2(6):563–7.

35. Zackowski KM, Dromerick AW, Sahrmann SA, Thach WT, Bastian AJ. How do strength, sensation, spasticity and joint individuation relate to the reaching deficits of people with chronic hemiparesis? Brain. 2004;127(5):1035–46.

36. Koh K, Oppizzi G, Kehs G, Zhang LQ. Abnormal coordination of upper extremity during target reaching in persons post stroke. Sci Rep. 2023;13(1):12838.

37. Paul T, Cieslak M, Hensel L, Wiemer VM, Tscherpel C, Grefkes C, et al. Corticospinal premotor fibers facilitate complex motor control after stroke. Ann Clin Transl Neurol. 2024 Sep;11(9):2439–49. doi:10.1002/acn3.52159

38. Unger RH, Lowe MJ, Beall EB, Bethoux F, Jones SE, Machado AG, et al. Stimulation of the Premotor Cortex Enhances Interhemispheric Functional Connectivity in Association with Upper Limb Motor Recovery in Moderate-to-Severe Chronic Stroke. Brain Connect. 2023 Oct;13(8):453–63. doi:10.1089/brain.2022.0064

39. Lin YL, Potter-Baker KA, Sankarasubramanian V, Cunningham DA, Li M, O’Laughlin K, et al. Stratification algorithm for repetitive TMS in stroke (START): Results from an exploratory crossover study. J Neurol Sci. 2025;473:123478.

40. Mustin M, Hensel L, Fink GR, Grefkes C, Tscherpel C. Individual contralesional recruitment in the context of structural reserve in early motor reorganization after stroke. NeuroImage. 2024;300:120828.

41. Casarotto A, Dolfini E, Russo M, Koch G, Fadiga L, D’Ausilio A. Superficial Ventral Premotor Pathways to Primary Motor Cortex Shape the Temporal Coordination of Precision Grasping. Eur J Neurosci. 2026 Jul;64(1):e70597. doi:10.1111/ejn.70597

42. Turrini S, Amadori A, Zago S, Arcara G, Avenanti A. Timing and circuit engagement shape state-dependent plasticity in the ventral premotor-motor pathway. Clin Neurophysiol. 2026;191:2112352. doi:10.1016/j.clinph.2026.2112352.

43. Vesia M, Prime SL, Yan X, Sergio LE, Crawford JD. Specificity of human parietal saccade and reach regions during transcranial magnetic stimulation. J Neurosci. 2010;30(39):13053–65.

44. Vesia M, Bolton DA, Mochizuki G, Staines WR. Human parietal and primary motor cortical interactions are selectively modulated during the transport and grip formation of goal-directed hand actions. Neuropsychologia. 2013;51(3):410–7.

45. Hensel L, Lange F, Tscherpel C, Viswanathan S, Freytag J, Volz LJ, et al. Recovered grasping performance after stroke depends on interhemispheric frontoparietal connectivity. Brain. 2023;146(3):1006–20.

46. Larivière S, Ward NS, Boudrias MH. Disrupted functional network integrity and flexibility after stroke: Relation to motor impairments. NeuroImage Clin. 2018;19:883–91.

47. Schaechter JD, Perdue KL. Enhanced cortical activation in the contralesional hemisphere of chronic stroke patients in response to motor skill challenge. Cereb Cortex. 2008;18(3):638–47.

48. Horn U, Roschka S, Eyme K, Walz AD, Platz T, Lotze M. Increased ventral premotor cortex recruitment after arm training in an fMRI study with subacute stroke patients. Behav Brain Res. 2016;308:152–9.

49. Allart E, Viard R, Lopes R, Devanne H, Delval A. Influence of motor deficiency and spatial neglect on the contralesional posterior parietal cortex functional and structural connectivity in stroke patients. Brain Topogr. 2020;33(2):176–90.

50. Zhang Y, Wang L, Yang J, Yan R, Zhang J, Sang L, et al. Abnormal functional networks in resting-state of the sub-cortical chronic stroke patients with hemiplegia. Brain Res. 2017;1663:51–8.

51. Lee J, Park E, Lee A, Chang WH, Kim DS, Kim YH. Alteration and role of interhemispheric and intrahemispheric connectivity in motor network after stroke. Brain Topogr. 2018;31(4):708–19.

52. Hordacre B, Lotze M, Jenkinson M, Lazari A, Barras CD, Boyd L, et al. Fronto-parietal involvement in chronic stroke motor performance when corticospinal tract integrity is compromised. NeuroImage Clin. 2021;29:102558.

53. Bestmann S, Swayne O, Blankenburg F, Ruff CC, Teo J, Weiskopf N, et al. The role of contralesional dorsal premotor cortex after stroke as studied with concurrent TMS-fMRI. J Neurosci. 2010;30(36):11926–37.

54. Goldenkoff ER, Deluisi JA, Destiny DP, Lee TG, Michon KJ, Brissenden JA, et al. The behavioral and neural effects of parietal theta burst stimulation on the grasp network are stronger during a grasping task than at rest. Front Neurosci. 2023;17:1198222.

55. Urbin MA, Tian J, Lafe CW, McKernan GP, Kortzorg N, Wyers L, et al. Perturbation of cortical activity elicits regional and age-dependent effects on unconstrained reaching behavior: a pilot study. Exp Brain Res. 2021 Dec;239(12):3585–600. doi:10.1007/s00221-021-06228-z

