## Supplementary Figure S1 for "Movement-specific effects of transcranial magnetic stimulation over premotor and parietal cortices after stroke"

### Supplementary materials

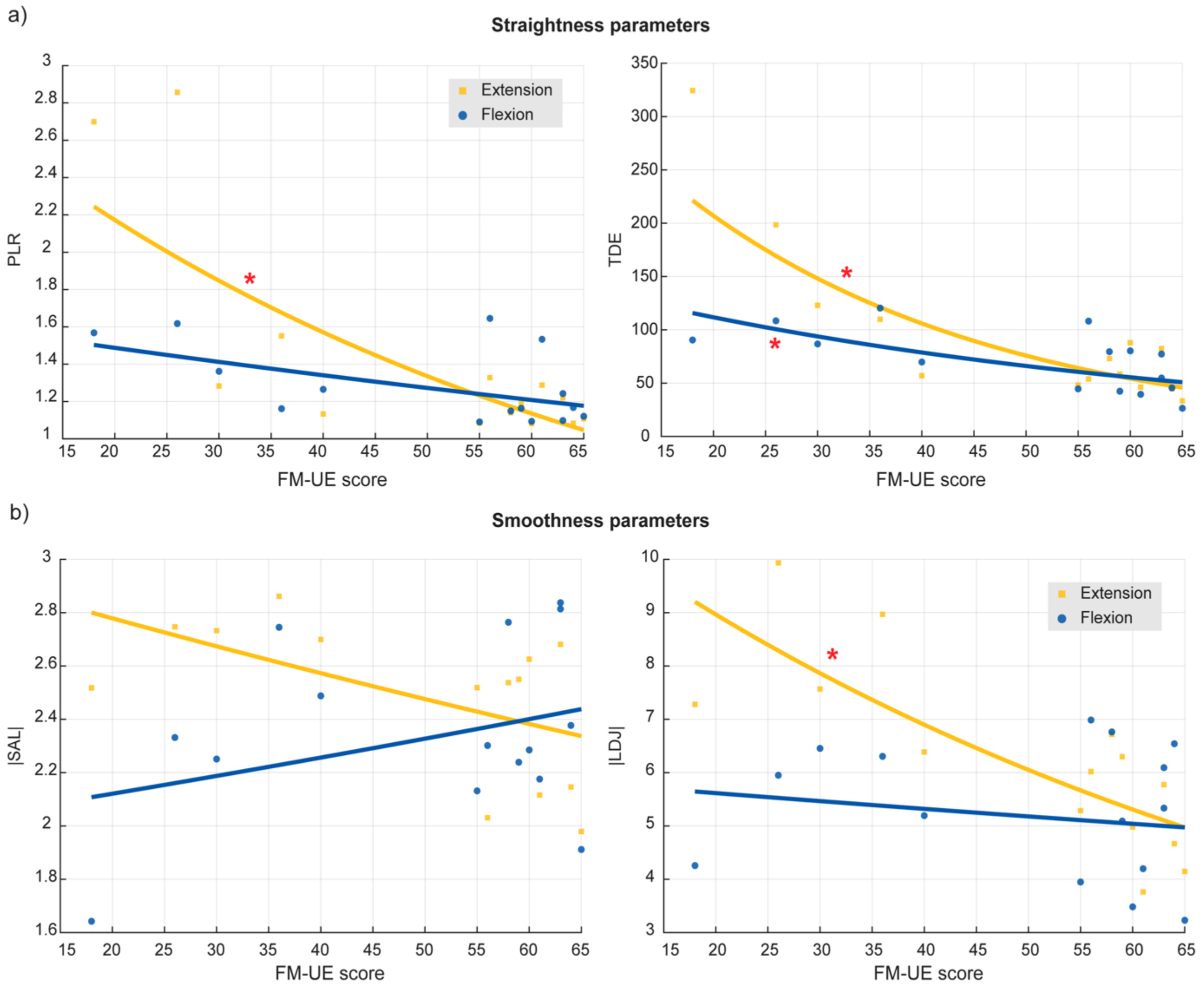


**Figure S1.** Relationship between FM-UE score and planar reaching performance in KINARM without stimulation. Participant-level mean values are shown for (a) trajectory straightness and (b) smoothness during extension and flexion. Lines represent model-estimated relationships with FM-UE, converted back to the original metric scale. Red asterisks indicate significant direction-specific associations after FDR correction. FM-UE, Fugl-Meyer Assessment of the Upper Extremity; PLR, path-to-length ratio; TDE, trajectory deviation error; |SAL|, absolute spectral arc length; |LDJ|, absolute log-dimensionless jerk.
